# ChatGPT versus human in generating medical graduate exam questions – An international prospective study

**DOI:** 10.1101/2023.05.13.23289943

**Authors:** Billy Ho Hung Cheung, Gary Kui Kai Lau, Gordon Tin Chun Wong, Elaine Yuen Phin Lee, Dhananjay Kulkarni, Choon Sheong Seow, Ruby Wong, Michael Tiong Hong Co

## Abstract

**Introduction:** This is a prospective study on the quality of multiple-choice questions (MCQs) generated by the language model ChatGPT for the use in medical graduate examination.

**Methods:** 50 MCQs were generated by ChatGPT with reference to two standard undergraduate medical textbooks (Harrison’s, and Bailey & Love’s). Another 50 MCQs were drafted by two university professoriate staffs using the same medical textbooks. All 100 MCQ were individually numbered, randomized and sent to five independent international assessors for MCQ quality assessment using a standardized assessment score on five assessment domains; namely, appropriateness of the question, clarity and specificity, relevance, discriminative power of alternatives, and suitability for medical graduate examination.

**Results:** The total time required for ChatGPT to create the 50 questions was 20 minutes 25 seconds while it took two human examiners a total of 211 minutes 33 seconds for drafting the 50 questions.

When a comparison of the mean score was made between the questions constructed by AI with those drafted by human, only in the relevance domain that the AI was inferior to human (AI: 7.56 +/- 0.94 vs human: 7.88 +/- 0.52; p = 0.04). There was no significant difference in question quality between questions drafted by AI versus human, in the total assessment score as well as in other domains.

Questions generated by AI yielded a wider range of scores while those created by human were consistent and within a narrower range.

**Conclusion:** ChatGPT has the potential to generate comparable-quality MCQs for medical graduate examination within a significantly shorter time.

## Introduction

The workload of university medical staff is a pressing issue that requires attention (Nassar et al., 2020). Medical staffs are often tasked with multiple responsibilities that can include, but not limit to, patient care, teaching, student assessment, research, and administrative works. This heavy workload can be especially challenging for medical academic staffs who are also required to uphold the standard of undergraduate medical exams (Epstein, 2007). The demand for exam quality has increased in recent years, as students and other stakeholders expect assessments to be fair, accurate, unbiased and align with the predefined learning objectives (Wong, Levinson, & Shojania, 2012; Yeoh, 2019).

In this context, the development of artificial intelligence (AI), machine learning, and language models offers a promising solution. AI is a rapidly developing field that has the potential to transform many industries, including education (Chen, Chen, & Lin, 2020). AI is a broad field that encompasses many different technologies, such as machine learning, natural language processing, and computer vision (Winston, 1992). Machine learning is a subset of AI that involves training algorithms to make predictions or decisions based on the database (Mitchell, 2007). This technology has been used in a variety of applications, including speech recognition, image classification, and language generation.

A large language model is a type of AI that has been trained on a massive amount of data and can generate human-like text with impressive accuracy (Brants, Popat, Xu, Och, & Dean, 2007). These models are called “large” because they have a huge number of parameters, which are the weights in the model’s mathematical equations that determine its behaviour. The more parameters a model has, the more information it can store and the more complex tasks it can perform.

ChatGPT is a specific type of large language model developed by OpenAI (AI, 2023). It is a state-of-the-art model that has been trained on a diverse range of texts, including news articles, books, and websites. This makes it highly versatile and capable of generating text on a wide range of topics with remarkable coherence and consistency. It is a pre-trained language model with knowledge up to 2021. However, it is possible to feed in relevant reference text by the operator, such that updated text or desired text outputs can be generated based on the operator’s command and preference.

The potential implications and possibilities of using ChatGPT for assessment in medical education are significant. A recent publication confirmed that the knowledge provided by ChatGPT is adequate to pass the United States Medical Licensing Exam (Kung et al., 2023). It is also believed that ChatGPT could be used to generate high-quality exam questions, provide personalized feedbacks to students, and to automate the grading process; hence reducing the workload of medical staffs and improving the quality of assessments (O’Connor & ChatGpt, 2023). This technology has the potential to be the game-changer in the field of medical education, providing new and innovative ways to assess student learning and evaluate exam quality.

Meanwhile, multiple-choice questions (MCQs) have been used as a form of knowledge-based assessment since the early twentieth century (Thomas M. Haladyna, Downing, & Rodriguez, 2002). MCQ is an important and integral component in both undergraduate and post-graduate exams, due to its standardization, equitability, objectiveness, cost effectiveness, and reliability (Kilgour & Tayyaba, 2016). When compared to essay type of questions or short answer questions, MCQs also allow assessment of a broader range of content – each exam paper can include large numbers of MCQs (Dion, St-Onge, Bartman, Touchie, & Pugh, 2022). This makes the MCQ format particularly suitable for summative final examinations. The major drawback to the MCQ format, however, is that high quality questions are difficult and time-consuming to draft.

Here, with an updated large language model AI available, we aim to evaluate the quality of exam questions generated by ChatGPT versus those drafted by university professoriate staffs based on international gold-standard medical reference textbooks.

## Methods

This is a prospective study to compare the quality of MCQs generated by ChatGPT versus those drafted by experienced university professoriate staffs for medical exam (Figure 1). The study is conducted in February 2023. To allow a fair comparison, certain criteria are set for question developments.

1. The questions were designed to meet standard for a medical graduate exam.
2. Only four choices were allowed for each question.
3. The questions were limited to knowledge-based questions only.
4. The questions were text-based only.
5. Topics regarding the exam context was set by an independent researcher before the design of the questions.
6. Both the professorial staff who designed the questions and the research operating ChatGPT were not allowed to view questions from the other side.
7. Standardized, internationally-used textbooks, with Harrison’s Principles of Internal Medicine 21th edition for medicine (Loscalzo et al., 2022), and Baily and Love’s Short Practice of Surgery 27th Edition for surgery (Williams, O’Connell, & McCaskie, 2018), were used as the reference for question generation.
8. Distractor were allowed to refer from sources other than the original reference provided.
9. No explanation was required for the question.

**Figure 1.**
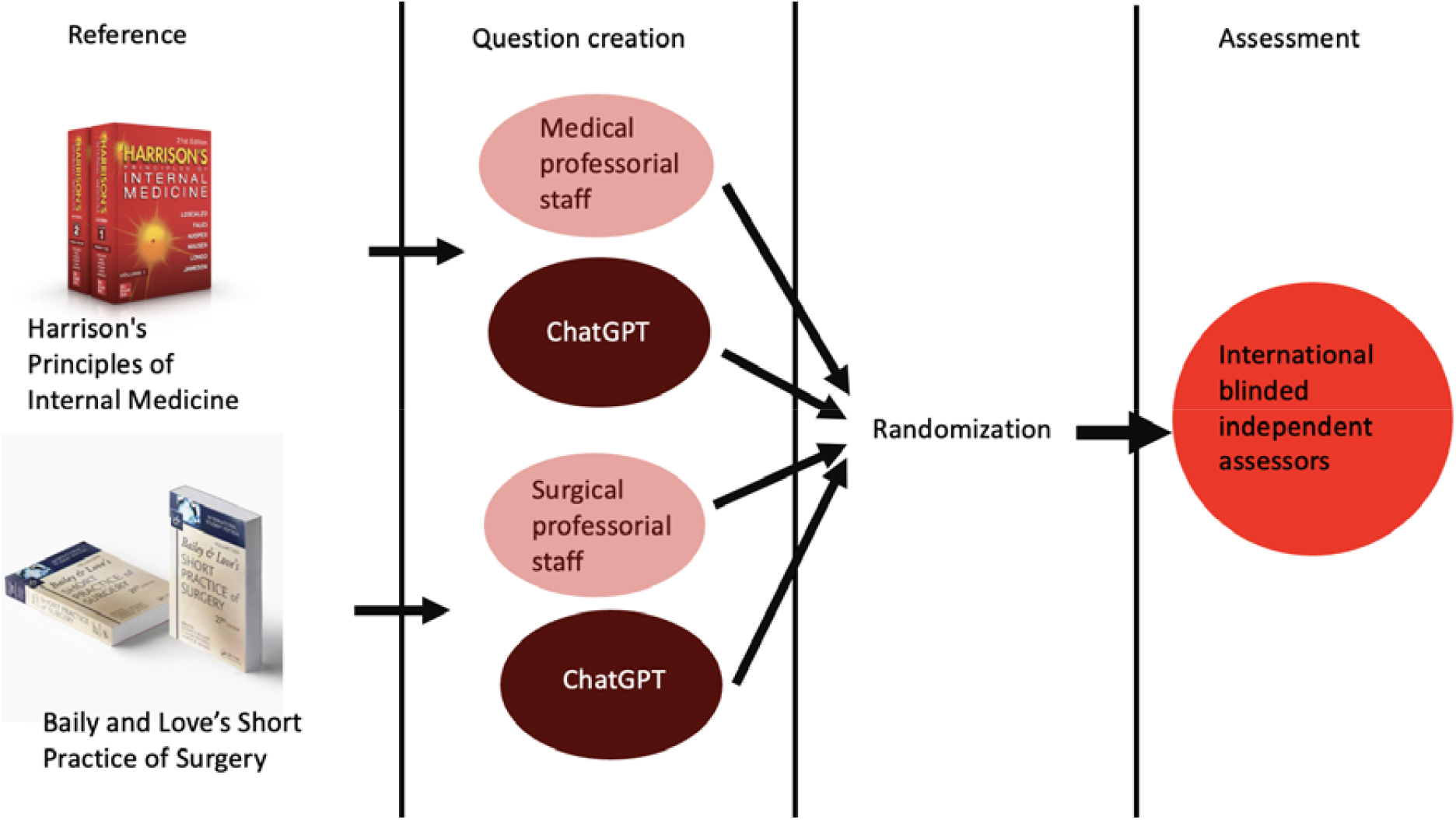
Schematic diagram of the study design.

ChatGPT plus (OpenAI, 2023) was used for question creation. This version is a small update to the standard ChatGPT version on 1sth of February 2023. According to the official webpage, ChatGPT plus carries the ability to answer follow-up questions, and challenge incorrect assumptions. This also provides better assess during peak hours, faster response time and priority to updates. To reduce memory retention bias, a new chat session was started in ChatGPT for each entry. The commander using the ChatGPT will only copy and paste the instruction to and the reference text to yield a question. Interaction is only allowed for clarification but not any modification. Questions provided by ChatGPT will be directly copied and used as the output for assessment by the independent quality assessment team.

### Question construction

50 multiple choice questions (MCQs) with four options covering Internal Medicine and Surgery were generated by the ChatGPT with reference from the two reference medical textbooks. Another 50 MCQs with four options were drafted by two experienced university professoriate staffs (with more than 15 years of clinical practice and academic experience in Internal Medicine and Surgery respectively) based on the same textbooks. Duration of work by AI was defined by the response time that ChatGPT needed to generate the questions once the prompt and the material was given and the time for the operator to copy and paste the questions were not included. The duration of work by ChatGPT and human were documented and compared.

### Blinded assessment by international expert panel

All 100 MCQs were individually numbered and randomized using computer-generated sequence.

A multiple-choice item consists of the stem, the options, and any auxiliary information (Vegada, Shukla, Khilnani, Charan, & Desai, 2016). The stem contains context, content, and/or the question the student is required to answer. The options include a set of alternative answers with one correct option and one or more incorrect options or distractors (Gierl, Bulut, Guo, & Zhang, 2017). Auxiliary information includes any additional content, in either the stem or option, required to generate an item. The incorrect options to a MCQ are known as distractors; they serve the purpose of diverting non-competent candidates away from the correct answer (Burton, Sudweeks, Merrill, & Wood, 1990), which they serve as an important hallmark of a high quality question (Lowe, 1991).

The question set were then sent to five independent international blinded assessors (From United Kingdom, Ireland, Singapore and Hong Kong) for assessment of the question quality. Members of the international panel of assessors are experienced clinicians with heavy involvement in medical education in their locality.

Quality of MCQs were objectively assessed by a numeric scale of 0 – 10 based on five assessment criteria. These include (I) Appropriateness of the question, defined as if the question is correct and appropriately constructed; (II) Clarity and specificity, defined as if the question is clear and specific without ambiguity; (III) Relevance, defined as the relevance to clinical context; (IV) Quality of the alternatives & discriminative power for the assessment of the choices provided; (V) and Suitability for graduate medical school exam. The total score of this section ranges from 0 to 50.

In addition, the assessors were also asked to determine if the questions were constructed by AI or by human, in which they were blinded about the total number of questions created by each arm. In addition, GPT-2 Output Detector, an AI output detector (OpenAI, 2022) was also used to predict as if the question was written by AI or human. This detector assigns a score between 0.02 and 99.98% to each question, with a higher score indicating a greater likelihood that the question was constructed by AI.

### Statistical analysis

All data were prospectively collected by a research assistant and computerized into a database. All statistical analysis were performed with the Statistical Product and Service Solution (SPSS) version 29. Comparison was made between questions created by AI and by human. Student T test or Mann-Whitney U test were used for comparison continuous variables for the five domains individually and combined. P value of less than 0.05 were considered statistically significant. Chi-squared test or Fisher’s exact test were used to compare discrete variables, namely the perception of the assessor whether a question was produced by AI or human.

A paired t-test was performed to assess for systematic differences between the mean measures of each rater (including GPT-2 Output Detector).

## Results

### Question construction

The question writing was performed by ChatGPT on two separate dates, 11^th^ and 17^th^ February. The work was carried out with stable internet via Wifi at a minimal of 15.40 Mbps for downloads and 11.96 Mbps for update speed.

The total time required for ChatGPT to create the 50 questions were 20 minutes 25 seconds while it took two human examiners a total of 211 minutes 33 seconds (84 minutes 56 seconds for the surgical examiner and 126 minutes 37 seconds for the medical examiner).

### Assessment of question quality

The results of the assessment by the independent blinded assessors were summarized in Table 1. We can see that the overall score was satisfactory with mean score of each domain above 7.

**Table 1.** Mean score of each domain.

| | Mean (± SD) | Range |
| --- | --- | --- |
| Appropriateness of the question | 7.78 ± 0.74 | 5.40 – 9.80 |
| Clarity and specificity | 7.63 ± 0.69 | 5.60 – 9.20 |
| Relevance | 7.72 ± 0.77 | 5.60 – 9.20 |
| Quality of the alternatives & discriminative power | 7.31 ± 0.65 | 5.60 – 7.31 |
| Suitability for graduate medical school exam | 7.32 ± 0.84 | 5.00 – 9.20 |
| Total score | 37.76 ± 3.34 | 27.20 – 46.20 |

When a comparison of the mean score was made between the questions constructed by AI with those constructed by human, only in the relevance category that the AI was inferior to human (AI: 7.56 ± 0.94 vs human: 7.88 ± 0.52; p = 0.04, Table 2). There was no significant difference in other domains of question quality assessment. The same applies to the total scores between AI and human (Table 1).

**Table 2.** Comparison of mean scores between questions generated by AI and human.

| | AI (± SD) | Human (± SD) | *P* |
| --- | --- | --- | --- |
| Appropriateness of the question | 7.72 ± 0.83 | 7.84 ± 0.65 | 0.45 |
| Clarity and specificity | 7.56 ± 0.81 | 7.69 ± 0.55 | 0.34 |
| Relevance | 7.56 ± 0.94 | 7.88 ± 0.52 | 0.04 |
| Quality of the alternatives & discriminative power | 7.26 ± 0.68 | 7.36 ± 0.61 | 0.46 |
| Suitability for graduate medical school exam | 7.25 ± 0.94 | 7.40 ± 0.72 | 0.39 |
| Total score | 37.36 ± 3.92 | 38.16 ± 2.62 | 0.23 |

SD = standard deviation

Questions generated by AI yielded a wider range of scores while those created by human were consistent and within a narrower range (Figure 2). Similar distribution was also observed across all five domains (Figure 3).

**Figure 2.**
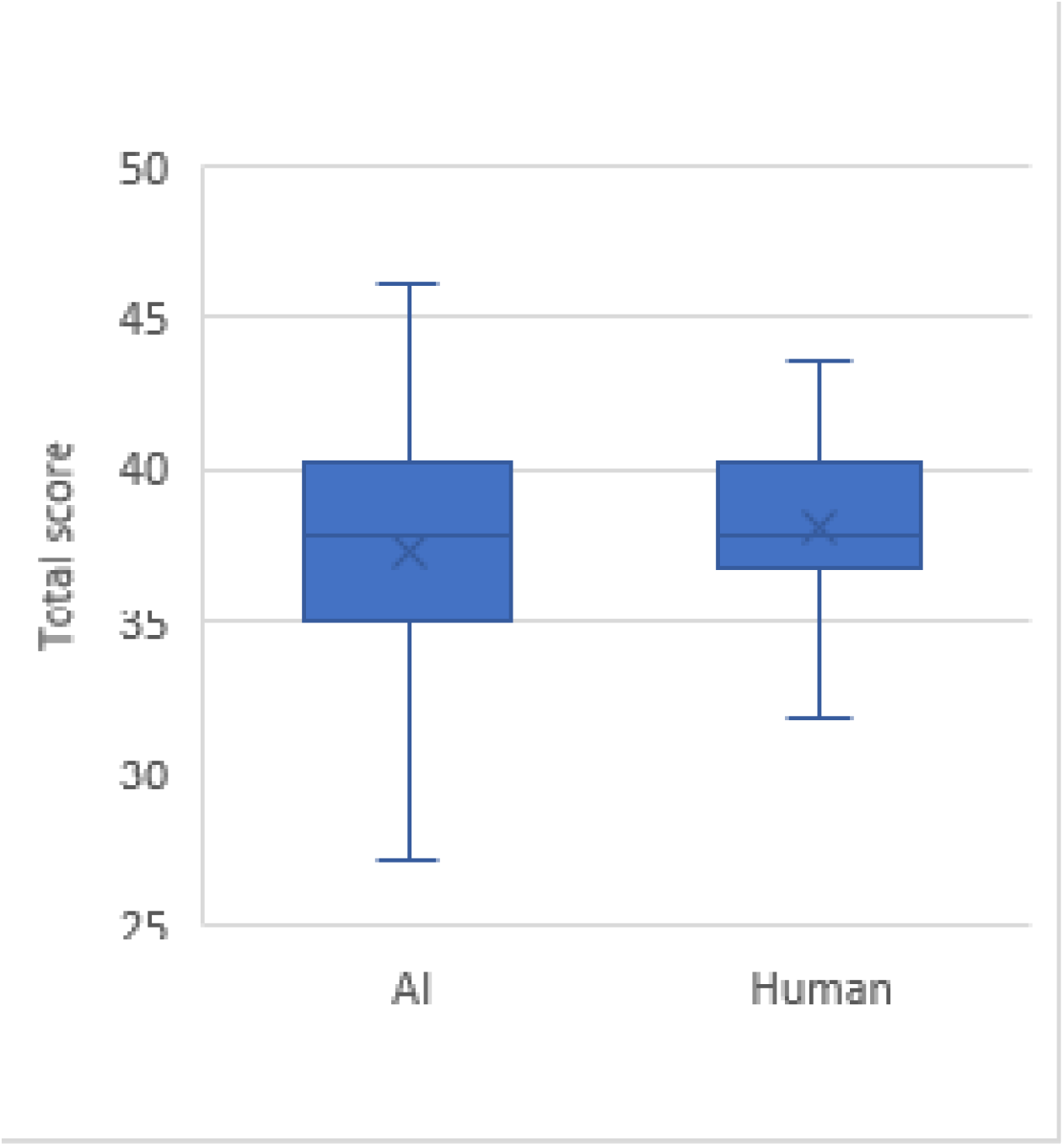
Assessment scores of MCQ quality between AI and human.

**Figure 3.**
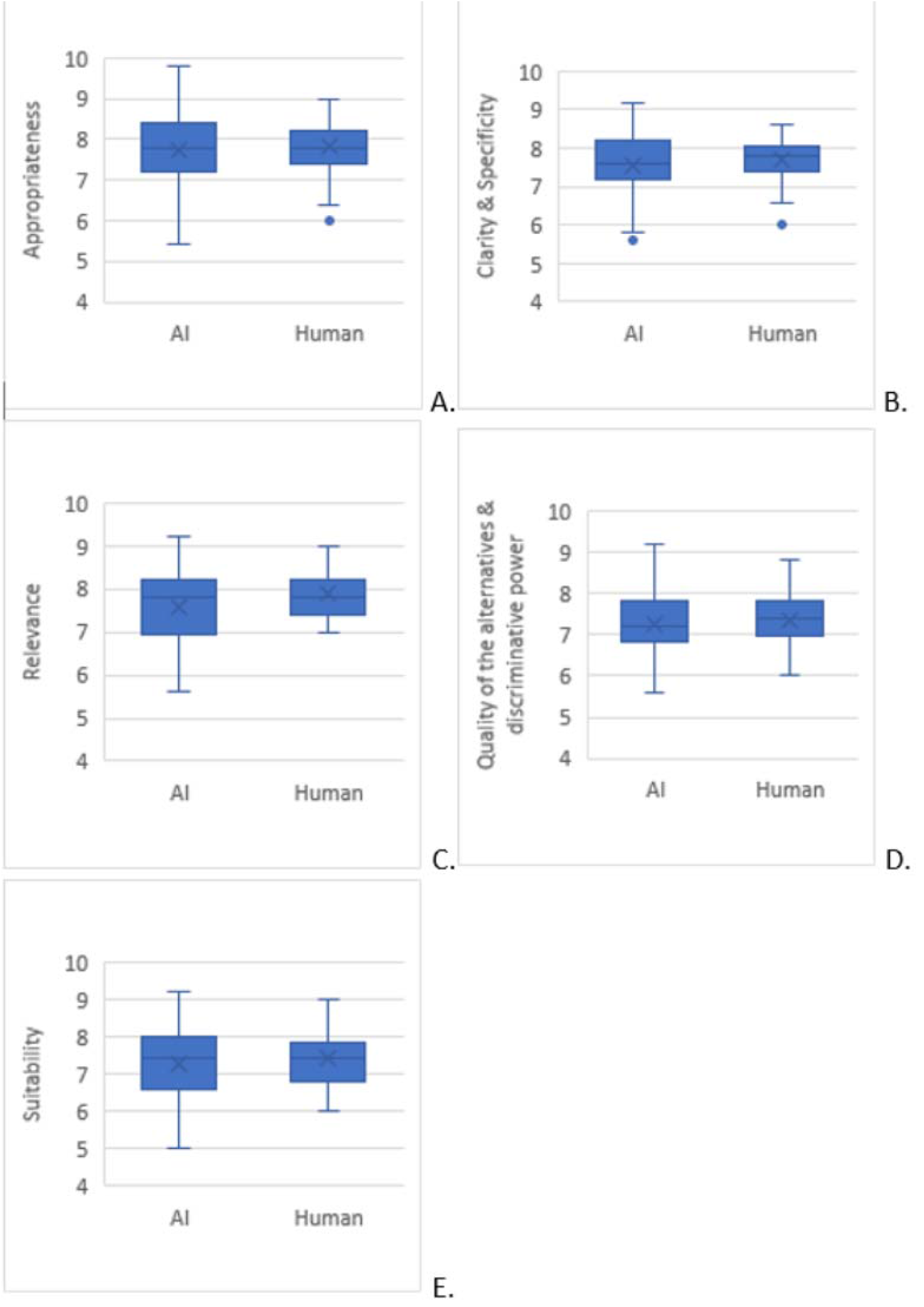
Assessment scores across all five assessment domains.

### AI vs human

When the questions generated by AI and human from the same reference textbooks were compared head-to-head, we can still see that questions generated by human generally received a higher mark, when compared to AI counterpart (Table 3). However, some questions drafted by AI surpassed human, from 36% to 44% across five assessment domains and including the total score.

**Table 3.**
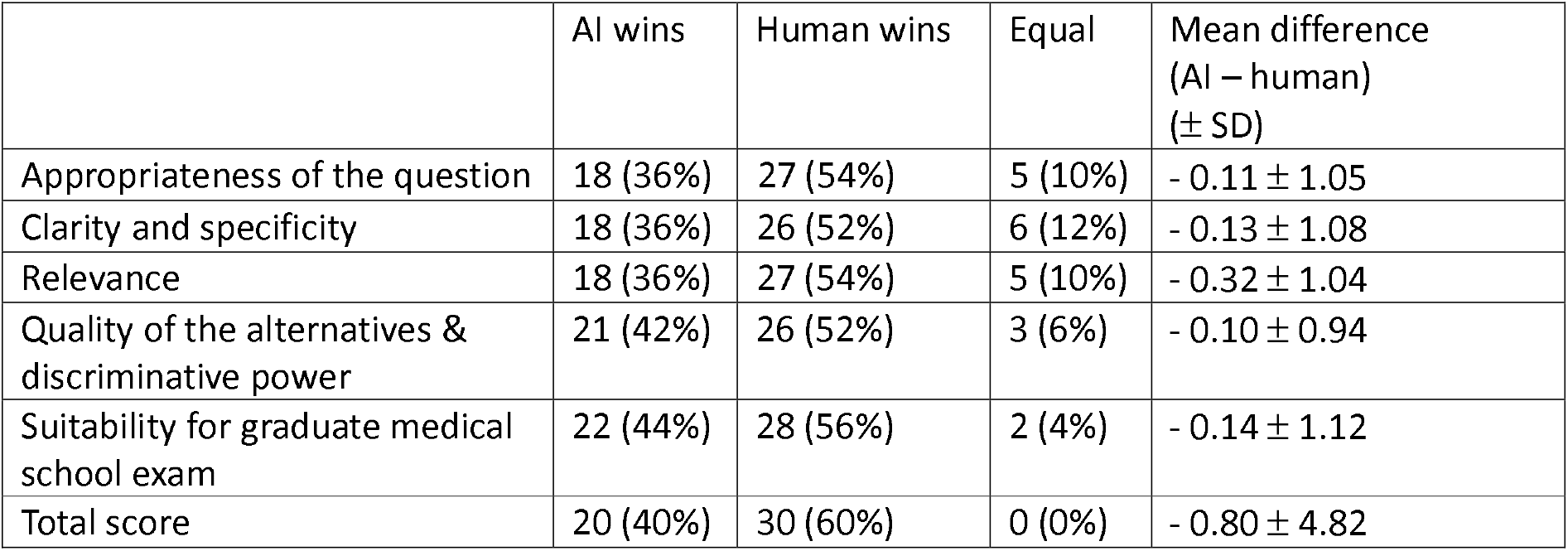
Comparison between AI vs human with the same reference.

### Blinded guess of the question writer by a panel of assessors

Assessors were asked to deduce if the question was written by AI or human and the results was shown in table 4. The results of assessment by GPT-2 Output Detector were also shown. The percentage of correct guess was consistently low. None of the five assessors, as well as the AI detector itself, could achieve a correct “guess” rate of 65% or above. In addition, our results showed that there is no correlation between the guess made by the assessor and the actual writer of the question.

**Table 4.** Blinded guess of question writer (i.e. AI vs Human)

|  | AI<br>(Correct guess, %) | Human<br>(Correct guess, %) | Correlation | p |
| --- | --- | --- | --- | --- |
| Assessor A | 51 (24, 48.0%) | 49 (23, 46.0%) | - 0.14 – 0.26 | 0.55 |
| Assessor B | 23 (14, 60.9%) | 77 (41, 53.2%) | - 0.38 – 0.10 | 0.24 |
| Assessor C | 59 (33, 55.9%) | 41 (24, 58.5%) | - 0.35 – 0.06 | 0.16 |
| Assessor D | 51 (27, 52.9%) | 49 (26, 53.1%) | - 0.26 – 0.14 | 0.55 |
| Assessor E | 44 (26, 59.1%) | 56 (32, 64.0%) | - 0.36 – 0.04 | 0.11 |
| GPT-2 Output<br>Detector | 12 (7, 58.3%) | 88 (45, 51.1%) | - 0.40 – 0.21 | 0.54 |

## Discussion

This is the first evidence in the literature showing that a commercially available, pre-trained AI can prepare exam material with a compatible quality to experienced human examiners.

MCQ is a crucial tool for assessment in education because they permit the direct measurement of many knowledge, skills, and competencies across a broad range of disciplines with the ability to test their concepts and principles, make judgments, drawing inferences, reasoning, interpretation of data, and information application (Kelly, 1916) (Dion et al., 2022; Thomas M. Haladyna et al., 2002). MCQs are also efficient to administer, easy to score objectively, and provide statistical information regarding the class performance on a particular question and to assess if the question was appropriate to the context that was presented (Thomas M Haladyna, 2004) (Iñarrairaegui et al., 2022). A standard MCQ consists of the stem, the options, and occasionally auxiliary information (Vegada et al., 2016). The stem contains context, content, and sets the question. The options include a set of alternative answers with one correct option and other incorrect options known as distractors (Gierl et al., 2017). Distractors are required to divert non-competent candidates away from the correct answer (Burton et al., 1990), which they serve as an important hallmark of a high quality question (Lowe, 1991). However, the major drawback to the MCQ format is that high quality questions are difficult, time-consuming and costly to write (Epstein, 2007). From our results, it is evident that even experienced examiners required on, average, more than ten minutes to prepare one question. With encouraging results demonstrated by the current study, we believe that AI could have to potential to generate quality MCQs for medical education.

Artificial Intelligence (AI) has been used in education to improve learning and teaching outcomes for several years. The history of AI in education can be traced back to the 1950s when scientists and mathematicians explored the mathematical possibility of AI (Haenlein & Kaplan, 2019). In recent years, AI has been used to analyze student learning abilities and history, allowing teachers to create the best learning program for all students (Chen et al., 2020).

The introduction of ChatGPT, an AI-powered chatbot, has also transformed the landscape of AI in education. ChatGPT was trained on a large dataset of real human conversations and online data, leading to its capability of song or poem writing, storytelling, list creation, and even passing exams (Kung et al., 2023).

However, the use of ChatGPT and other AI-powered tools in education has also raised concerns about their negative impacts on student learning (O’Connor & ChatGpt, 2023). Including ChatGPT, most AI models are trained by the vast content available on the internet, which their reliability and credibility are questionable. Moreover, many AIs were found to have significant bias due to their training data (Howard et al., 2021). Another major potential setback related to natural language generator AI is called hallucination (Maynez, Narayan, Bohnet, & McDonald, 2020). Like hallucination described in human, this condition refers to a phenomenon where the AI generates nonsensical, or unfaithful to the provided source input. This has led to immediate recall even for some initially promising AI from some of the largest internet companies, such as Galactica from Meta Inc (Heaven, 2023). Hence, our team proposed the use of the AI by the educators, which demonstrate a feasible way of utilization for the educational purpose. To minimize bias and hallucination, our proposed methodology consists of providing reliable reference for the AI to generate questions instead of complete dependence on its own database <u>(Alkaissi & McFarlane, 2023).</u>.

When the questions generated by ChatGPT were reviewed, we could also observe that they were compatible with the guidance from the Division of Education, American College of Surgeons, with minimal negative features, including a minimal use of negative stem (only 14% (7/50), compared to 12% (6/50) by human examiners) with a lack of “except”, “All/none of the above” (Brame, 2013). However, we also acknowledge that there were intrinsic limitations with ChatGPT in generating MCQs for medial graduate examinations. First, as it is a pure language-based model, it cannot generate any text and correlate it with clinical photo of radiological images, which is also an important area in the exam assessing candidates’ interpretation skills. In addition, our pre-study tests have found that ChatGPT performed poorly when it was instructed to generate a clinical scenario, possibly due to the high complexity of knowledge and experience required to create a relevant scenario, limiting its use to assess candidates’ ability of application of their knowledge. However, with the continuous effort from the OpenAI team and the rapid evolution of AI technology, these barriers might be solved in the near future (OpenAI, 2012).

## Limitations

The first limitation is that the reference material used was obtained directly from a textbook, and the length of the text was limited by the AI platform which is currently at 510 tokens, potentially leading to selection bias by the operator. In contrast, human exam writers can recall information associated with the text, resulting in higher quality questions. Another limitation is that only two professoriate staffs from Medicine and Surgery departments, but not experts in all fields, generated the MCQs in this study. This is different from the real-life scenario where graduate exam questions were generated by a large pool of question writers from different clinical departments, which were then reviewed and vetted by a panel of professoriate staffs. The third limitation is related to the absence of human interference in the question writing process. AI generated the questions, and the first-available question was captured without any polishing, while ChatGPT is also known to be sensitive to small changes in the prompt and can provide various answers even with the same prompt. In addition, this study only evaluated the use of AI in generating MCQs, full application of AI in generating the entire set of medical exam questions is yet to be fully evaluated. Lastly, with the improvements in newer generation ChatGPT AI platform and other adjuncts. AI may perform better than what was observed in this current study. Nonetheless, this study provided solid evidence of the ability of ChatGPT and its strong potential in assisting medical exam MCQ preparation.

## Conclusion

This is the first study showing that ChatGPT, an open-sourced AI platform, can be utilized as an exam question writer for graduate medical exam with a comparable performance to experienced human examiners. Our study supports the continuous exploration of how large language model AI can assist the academia to improve their efficiency while maintaining a consistently high standard. Further studies are required to further explore applications and other limitations of the booming AI platforms to improve reliability with minimal bias.

## Data Availability

All data produced in the present study are available upon reasonable request to the authors
All data produced in the present work are contained in the manuscript

## Notes

### Competing Interest Statement

The authors have declared no competing interest.

### Funding Statement

This study did not receive any funding

